# The Impact of Antihypertensive De-escalation: A Secondary Analysis of SPRINT

**DOI:** 10.1101/2025.04.17.25326047

**Authors:** Fanxing Du, Steven M. Smith, Mark S. Segal, Adam P. Bress, Tianze Jiao

## Abstract

**Background:** Antihypertensive de-escalation—reducing the number of antihypertensive medications—is sometimes warranted but may increase cardiovascular risk if not clinically appropriate. Our study investigated the impact of antihypertensive de-escalation without clinical consideration on cardiovascular outcomes and adverse events.

**Method:** This is a secondary analysis of Systolic Blood Pressure Intervention Trial (SPRINT), a randomized open-label trial, compared intensive (target systolic blood pressure (SBP) <120mmHg) vs. standard (target SBP <140mmHg) antihypertensive strategies. Before randomization, some patients underwent antihypertensive de-escalation to increase SBP into 130-180mmHg for eligibility. The exposure was de-escalation versus control, measured by the change in the number of antihypertensive medications between the screening visit and randomization. The primary outcome was major cardiovascular events (MACE), comprising myocardial infarction, acute coronary syndrome, stroke, heart failure, or cardiovascular cause death. Secondary outcomes included myocardial infarction, stroke, heart failure, and all-cause mortality. Adverse events include hypotension, syncope, acute kidney injury, bradycardia, and electrolyte abnormalities.

**Results:** The number of patients in de-escalation and control groups were 427 vs. 4,007 in the intensive arm, and 794 vs. 3,733 in the standard arm. De-escalation patients had higher baseline cardiovascular risk. In the standard arm, de-escalation was associated with increased MACE risk (HR 1.34, 95% CI: 1.04–1.74). In contrast, no excess MACE risk was observed in the intensive arm (SBP <120 mmHg).

**Conclusion:** Antihypertensive de-escalation without clinical consideration is associated with an increased risk of MACE. It reassures the importance of intensive SBP on patients at high cardiovascular risk and indicates a more intensive SBP target may be needed.

## Introduction

Hypertension is highly prevalent among older people^1^, and is recognized as a leading risk factor for major cardiovascular diseases and all-cause death globally^2,3^. Previous studies showed a U-shape association between blood pressure (BP) and major adverse cardiovascular events^4–7^. Thus, an optimal systolic blood pressure (SBP) range may exist where the best clinical benefit could be achieved^8^. However, the recommended target BP varies between different guidelines issued by various institutions over the years. For example, World Health Organization (WHO) 2021^9^, American Heart Association (AHA) 2020^10^ and UK National Institute for Health and Care Excellence (NICE) 2019^11^ guidelines suggest an SBP target <140 mmHg, while European Society of Hypertension (ESH)/European Society of Cardiology (ESC), American Society of Hypertension (ASH), and U.S. guidance from the Joint National Committee (JNC) 8 in the USA between 2011 and 2015 recommended a target ranging from <130 to <150 mmHg^12^. Even the recommendations from JNC 8 were controversial, with some authors supporting a target of <150 mmHg^13^, while others advocated for <140 mmHg^14^. Besides, the target for certain populations also varies: patients with chronic kidney disease or diabetes usually have lower recommended SBP targets, e.g., <130 mmHg^11^.

In clinical practice, physicians may set individualized SBP targets based on local guidelines, clinical experience, and patient preference^15^. When the target SBP increases, or patients experience episodes of low SBP, physicians may de-escalate the number of antihypertensive medications patients receive. Typically, de-escalation is triggered by clinical factors such as improved comorbidities^16^ and concerns about adverse events or drug-drug interactions^17^.

De-escalation, when clinically appropriate, is crucial to avoid polypharmacy among the elderly, which are associated with an increasing risk of death^18^. However, de-escalation sometimes relates to non-clinical factors, such as the healthcare providers’ preferences^19^, patients’ poor adherence to medications^20^, limited access to medications^21^, and affordability issues^22^. De-escalations caused by these factors may lead to harm for certain patient populations who may warrant a lower target BP. Yet, this association has not been properly studied.

Recently, there has been an increasing interest in de-escalation of antihypertensives, especially for elderly, given the concerns of adverse events, polypharmacy, drug-drug interaction and cognitive reduction. One type of de-escalation is the intentional discontinuation of a medication when it is no longer beneficial, which is also called deprescribing^23,24^. This kind of de-escalation may reduce the risk of adverse events such as acute kidney injury, hyperkalemia, hypotension, and syncope^25^. Besides, it can help reduce polypharmacy, minimize drug-drug interactions, and lower the risk of cognitive decline^26^. However, de-escalation without clinical consideration may adversely affect cardiovascular risk, and few studies have quantitatively assessed the cardiovascular risk associated with antihypertensive de-escalation due to non-clinical factors. Moreover, de-escalated patients may have different characteristics and antihypertensive treatment patterns from others due to the arbitrary relaxing of the target SBP. Thus, there is an urgent need to bridge these knowledge gaps to better understand the impact of de-escalation on cardiovascular events with real-world evidence.

Investigating de-escalation in the real-world setting is often challenging due to the absence of data on a target SBP, unknown reasons for de-escalation and irregular follow-up visits. Therefore, we sought to conduct a post-hoc analysis of the Systolic Blood Pressure Intervention Trial (SPRINT) which offers a unique scenario to study the impact of de-escalation, given that a significant number of SPRINT patients received treatment de-escalation solely to meet the trial’s inclusion criteria (SBP: 130-180 mmHg) before enrollment and were randomly assigned to a target SBP goal above their usual SBP. Specifically, our post hoc analysis of SPRINT data aimed to 1) investigate the impact of antihypertensive de-escalation on cardiovascular outcomes and adverse events; and 2) identify the characteristics of de-escalated patients.

## Methods

### Study design and population

Details about the design and implementation of SPRINT have been published elsewhere^5,7,10^. Briefly, SPRINT was a randomized, controlled, open-label trial that was conducted at 102 clinical sites in the United States from November 2010 to August 2015. Participants included adults aged ≥50 years with an SBP of 130 to 180 mmHg and a high risk of cardiovascular disease (CVD). Participants were randomly assigned to the intensive treatment arm (SBP target: <120 mmHg) or standard treatment arm (SBP target: <140 mmHg) and then received continuous antihypertensive treatment adjustment to achieve assigned SBP targets. Patients had routine, monthly visits for the first 3 months and every 3 months thereafter until end of follow-up (up to 5 years). The trial was stopped early (median follow-up time, 3.33 years) because of substantive evidence of treatment benefit^11^. The SPRINT was approved by the Institutional Review Board at each study site, with monitoring provided by an independent Data and Safety Monitoring Board^27^. This study was approved by the Institutional Review Boards of the University of Florida. We used the full SPRINT individual participant dataset.

### Exposure and Outcomes

Our primary exposure was treatment de-escalation, measured as a decrease in the number of antihypertensive medications prescribed from the screening visit to the randomization visit, regardless of the randomized intervention (target SBP <120 or 140 mmHg) assignment. Patients who received equal or more antihypertensive medications at randomization, compared to the screening visit, were assigned to the control group.

The primary outcome mirrored the original SPRINT trial primary outcome, i.e., a major adverse cardiovascular event (MACE), defined as a composite of the first occurrence of myocardial infarction, acute coronary syndrome (not resulting in myocardial infarction), stroke, acute decompensated heart failure, or death from cardiovascular causes. Secondary outcomes included myocardial infarction, stroke, heart failure, and all-cause mortality. Adverse events included hypotension, syncope, acute kidney injury, bradycardia, and electrolyte abnormalities^27^.

### Measurements

Baseline characteristics collected included age, sex, race, smoking status, alcohol use, total cholesterol, HDL cholesterol, total triglycerides, body mass index (BMI), serum glucose, estimated glomerular filtration rate (eGFR), Framingham 10-year cardiovascular disease risk score, comorbidities and medications use (Table 1). Patients’ SBP, diastolic blood pressure (DBP), and antihypertensive medication received were obtained at baseline and at every routine visit during follow-up. Antihypertensive medication received before randomization was also collected at the screening visit.

**Table 1.**
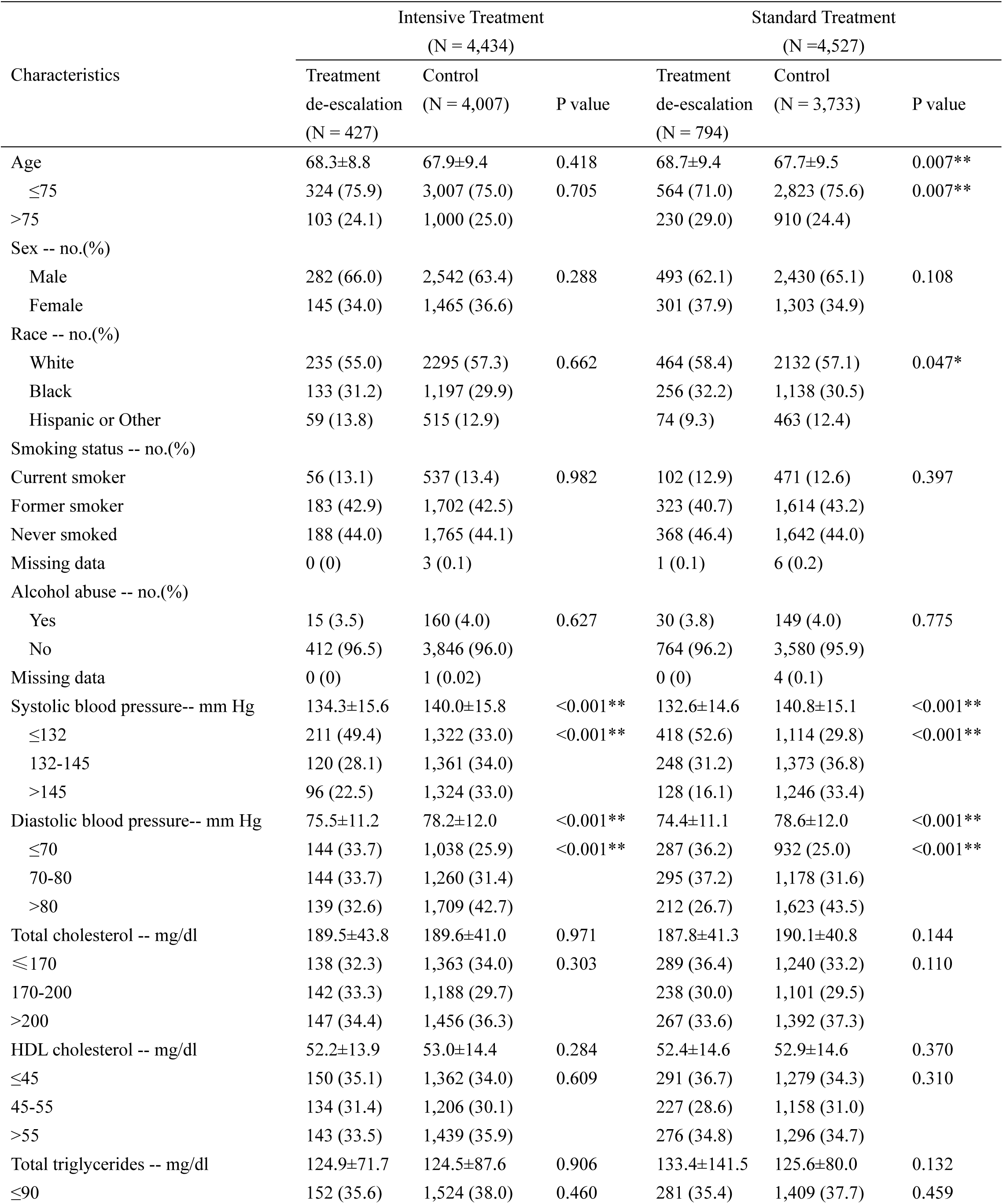
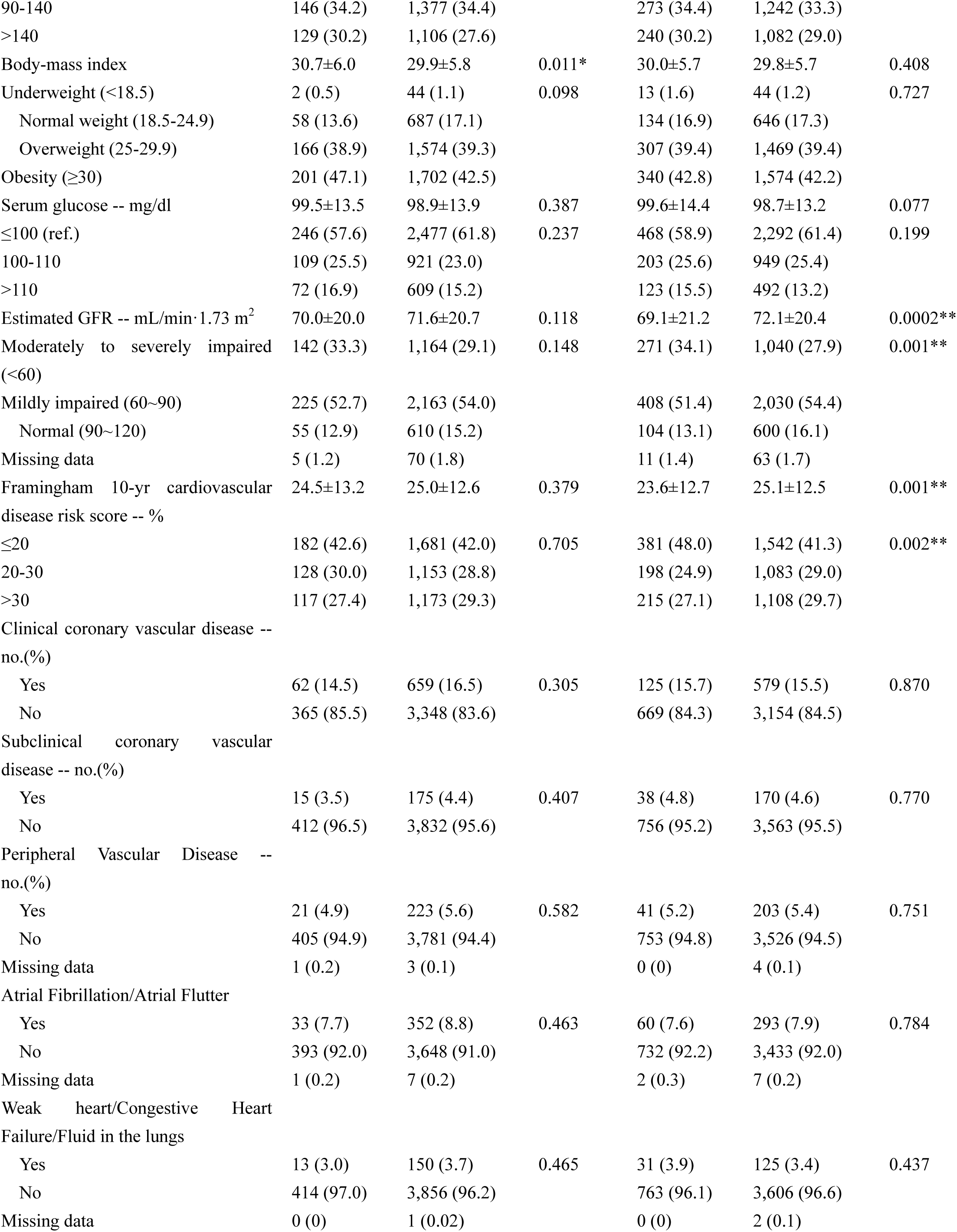
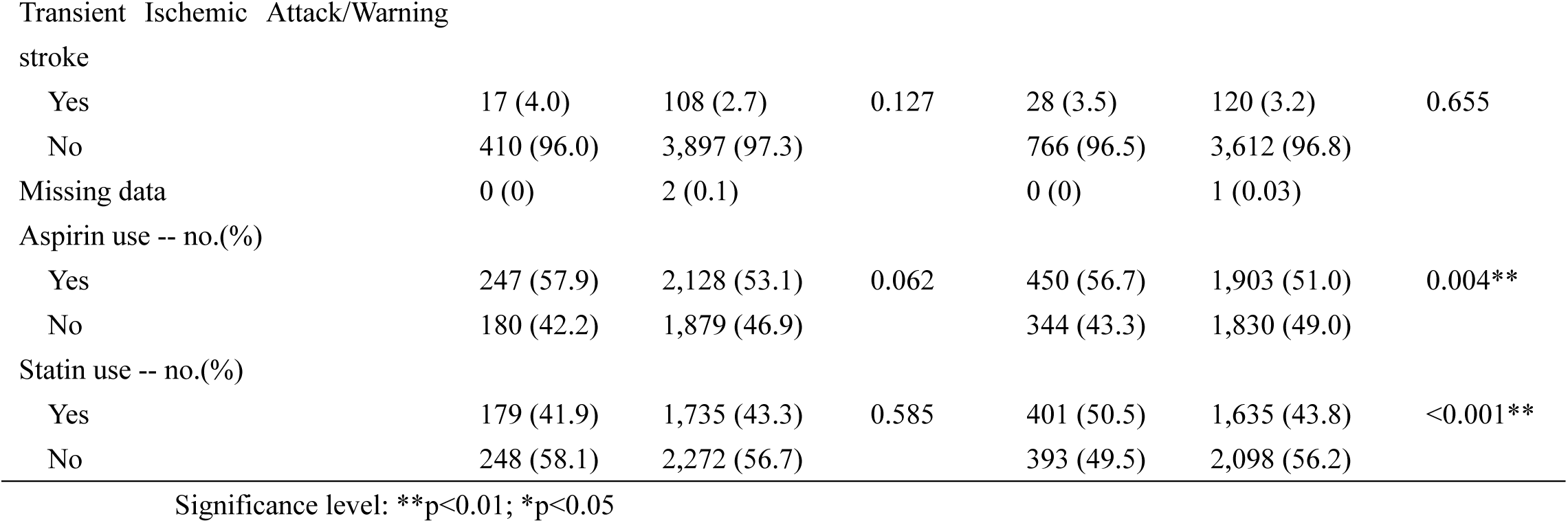
Baseline Characteristics of the Study Participants.

| Characteristics | Intensive Treatment<br>(N = 4,434) |  |  | Standard Treatment<br>(N = 4,527) |  |  |
| --- | --- | --- | --- | --- | --- | --- |
|  | Treatment | Control | P value | Treatment | Control | P value |
|  | de-escalation<br>(N = 427) | (N = 4,007) |  | de-escalation<br>(N = 794) | (N = 3,733) |  |
| Age | 68.3±8.8 | 67.9±9.4 | 0.418 | 68.7±9.4 | 67.7±9.5 | 0.007** |
| ≤75 | 324 (75.9) | 3,007 (75.0) | 0.705 | 564 (71.0) | 2,823 (75.6) | 0.007** |
| >75 | 103 (24.1) | 1,000 (25.0) |  | 230 (29.0) | 910 (24.4) |  |
| Sex -- no.(%) |  |  |  |  |  |  |
| Male | 282 (66.0) | 2,542 (63.4) | 0.288 | 493 (62.1) | 2,430 (65.1) | 0.108 |
| Female | 145 (34.0) | 1,465 (36.6) |  | 301 (37.9) | 1,303 (34.9) |  |
| Race -- no.(%) |  |  |  |  |  |  |
| White | 235 (55.0) | 2295 (57.3) | 0.662 | 464 (58.4) | 2132 (57.1) | 0.047* |
| Black | 133 (31.2) | 1,197 (29.9) |  | 256 (32.2) | 1,138 (30.5) |  |
| Hispanic or Other | 59 (13.8) | 515 (12.9) |  | 74 (9.3) | 463 (12.4) |  |
| Smoking status -- no.(%) |  |  |  |  |  |  |
| Current smoker | 56 (13.1) | 537 (13.4) | 0.982 | 102 (12.9) | 471 (12.6) | 0.397 |
| Former smoker | 183 (42.9) | 1,702 (42.5) |  | 323 (40.7) | 1,614 (43.2) |  |
| Never smoked | 188 (44.0) | 1,765 (44.1) |  | 368 (46.4) | 1,642 (44.0) |  |
| Missing data | 0 (0) | 3 (0.1) |  | 1 (0.1) | 6 (0.2) |  |
| Alcohol abuse -- no.(%) |  |  |  |  |  |  |
| Yes | 15 (3.5) | 160 (4.0) | 0.627 | 30 (3.8) | 149 (4.0) | 0.775 |
| No | 412 (96.5) | 3,846 (96.0) |  | 764 (96.2) | 3,580 (95.9) |  |
| Missing data | 0 (0) | 1 (0.02) |  | 0 (0) | 4 (0.1) |  |
| Systolic blood pressure-- mm Hg | 134.3±15.6 | 140.0±15.8 | <0.001** | 132.6±14.6 | 140.8±15.1 | <0.001** |
| ≤132 | 211 (49.4) | 1,322 (33.0) | <0.001** | 418 (52.6) | 1,114 (29.8) | <0.001** |
| 132-145 | 120 (28.1) | 1,361 (34.0) |  | 248 (31.2) | 1,373 (36.8) |  |
| >145 | 96 (22.5) | 1,324 (33.0) |  | 128 (16.1) | 1,246 (33.4) |  |
| Diastolic blood pressure-- mm Hg | 75.5±11.2 | 78.2±12.0 | <0.001** | 74.4±11.1 | 78.6±12.0 | <0.001** |
| ≤70 | 144 (33.7) | 1,038 (25.9) | <0.001** | 287 (36.2) | 932 (25.0) | <0.001** |
| 70-80 | 144 (33.7) | 1,260 (31.4) |  | 295 (37.2) | 1,178 (31.6) |  |
| >80 | 139 (32.6) | 1,709 (42.7) |  | 212 (26.7) | 1,623 (43.5) |  |
| Total cholesterol -- mg/dl | 189.5±43.8 | 189.6±41.0 | 0.971 | 187.8±41.3 | 190.1±40.8 | 0.144 |
| ≤170 | 138 (32.3) | 1,363 (34.0) | 0.303 | 289 (36.4) | 1,240 (33.2) | 0.110 |
| 170-200 | 142 (33.3) | 1,188 (29.7) |  | 238 (30.0) | 1,101 (29.5) |  |
| >200 | 147 (34.4) | 1,456 (36.3) |  | 267 (33.6) | 1,392 (37.3) |  |
| HDL cholesterol -- mg/dl | 52.2±13.9 | 53.0±14.4 | 0.284 | 52.4±14.6 | 52.9±14.6 | 0.370 |
| ≤45 | 150 (35.1) | 1,362 (34.0) | 0.609 | 291 (36.7) | 1,279 (34.3) | 0.310 |
| 45-55 | 134 (31.4) | 1,206 (30.1) |  | 227 (28.6) | 1,158 (31.0) |  |
| >55 | 143 (33.5) | 1,439 (35.9) |  | 276 (34.8) | 1,296 (34.7) |  |
| Total triglycerides -- mg/dl | 124.9±71.7 | 124.5±87.6 | 0.906 | 133.4±141.5 | 125.6±80.0 | 0.132 |
| ≤90 | 152 (35.6) | 1,524 (38.0) | 0.460 | 281 (35.4) | 1,409 (37.7) | 0.459 |
| 90-140 | 146 (34.2) | 1,377 (34.4) |  | 273 (34.4) | 1,242 (33.3) |  |
| >140 | 129 (30.2) | 1,106 (27.6) |  | 240 (30.2) | 1,082 (29.0) |  |
| Body-mass index | 30.7±6.0 | 29.9±5.8 | 0.011* | 30.0±5.7 | 29.8±5.7 | 0.408 |
| Underweight (<18.5) | 2 (0.5) | 44 (1.1) | 0.098 | 13 (1.6) | 44 (1.2) | 0.727 |
| Normal weight (18.5-24.9) | 58 (13.6) | 687 (17.1) |  | 134 (16.9) | 646 (17.3) |  |
| Overweight (25-29.9) | 166 (38.9) | 1,574 (39.3) |  | 307 (39.4) | 1,469 (39.4) |  |
| Obesity (≥30) | 201 (47.1) | 1,702 (42.5) |  | 340 (42.8) | 1,574 (42.2) |  |
| Serum glucose -- mg/dl | 99.5±13.5 | 98.9±13.9 | 0.387 | 99.6±14.4 | 98.7±13.2 | 0.077 |
| ≤100 (ref.) | 246 (57.6) | 2,477 (61.8) | 0.237 | 468 (58.9) | 2,292 (61.4) | 0.199 |
| 100-110 | 109 (25.5) | 921 (23.0) |  | 203 (25.6) | 949 (25.4) |  |
| >110 | 72 (16.9) | 609 (15.2) |  | 123 (15.5) | 492 (13.2) |  |
| Estimated GFR -- mL/min·1.73 m<sup>2</sup> | 70.0±20.0 | 71.6±20.7 | 0.118 | 69.1±21.2 | 72.1±20.4 | 0.0002** |
| Moderately to severely impaired (<60) | 142 (33.3) | 1,164 (29.1) | 0.148 | 271 (34.1) | 1,040 (27.9) | 0.001** |
| Mildly impaired (60~90) | 225 (52.7) | 2,163 (54.0) |  | 408 (51.4) | 2,030 (54.4) |  |
| Normal (90~120) | 55 (12.9) | 610 (15.2) |  | 104 (13.1) | 600 (16.1) |  |
| Missing data | 5 (1.2) | 70 (1.8) |  | 11 (1.4) | 63 (1.7) |  |
| Framingham 10-yr cardiovascular disease risk score -- % | 24.5±13.2 | 25.0±12.6 | 0.379 | 23.6±12.7 | 25.1±12.5 | 0.001** |
| ≤20 | 182 (42.6) | 1,681 (42.0) | 0.705 | 381 (48.0) | 1,542 (41.3) | 0.002** |
| 20-30 | 128 (30.0) | 1,153 (28.8) |  | 198 (24.9) | 1,083 (29.0) |  |
| >30 | 117 (27.4) | 1,173 (29.3) |  | 215 (27.1) | 1,108 (29.7) |  |
| Clinical coronary vascular disease -- no.(%) |  |  |  |  |  |  |
| Yes | 62 (14.5) | 659 (16.5) | 0.305 | 125 (15.7) | 579 (15.5) | 0.870 |
| No | 365 (85.5) | 3,348 (83.6) |  | 669 (84.3) | 3,154 (84.5) |  |
| Subclinical coronary vascular disease -- no.(%) |  |  |  |  |  |  |
| Yes | 15 (3.5) | 175 (4.4) | 0.407 | 38 (4.8) | 170 (4.6) | 0.770 |
| No | 412 (96.5) | 3,832 (95.6) |  | 756 (95.2) | 3,563 (95.5) |  |
| Peripheral Vascular Disease -- no.(%) |  |  |  |  |  |  |
| Yes | 21 (4.9) | 223 (5.6) | 0.582 | 41 (5.2) | 203 (5.4) | 0.751 |
| No | 405 (94.9) | 3,781 (94.4) |  | 753 (94.8) | 3,526 (94.5) |  |
| Missing data | 1 (0.2) | 3 (0.1) |  | 0 (0) | 4 (0.1) |  |
| Atrial Fibrillation/Atrial Flutter |  |  |  |  |  |  |
| Yes | 33 (7.7) | 352 (8.8) | 0.463 | 60 (7.6) | 293 (7.9) | 0.784 |
| No | 393 (92.0) | 3,648 (91.0) |  | 732 (92.2) | 3,433 (92.0) |  |
| Missing data | 1 (0.2) | 7 (0.2) |  | 2 (0.3) | 7 (0.2) |  |
| Weak heart/Congestive Heart Failure/Fluid in the lungs |  |  |  |  |  |  |
| Yes | 13 (3.0) | 150 (3.7) | 0.465 | 31 (3.9) | 125 (3.4) | 0.437 |
| No | 414 (97.0) | 3,856 (96.2) |  | 763 (96.1) | 3,606 (96.6) |  |
| Missing data | 0 (0) | 1 (0.02) |  | 0 (0) | 2 (0.1) |  |
| Transient Ischemic Attack/Warning stroke |  |  |  |  |  |  |
| Yes | 17 (4.0) | 108 (2.7) | 0.127 | 28 (3.5) | 120 (3.2) | 0.655 |
| No | 410 (96.0) | 3,897 (97.3) |  | 766 (96.5) | 3,612 (96.8) |  |
| Missing data | 0 (0) | 2 (0.1) |  | 0 (0) | 1 (0.03) |  |
| Aspirin use -- no.(%) |  |  |  |  |  |  |
| Yes | 247 (57.9) | 2,128 (53.1) | 0.062 | 450 (56.7) | 1,903 (51.0) | 0.004** |
| No | 180 (42.2) | 1,879 (46.9) |  | 344 (43.3) | 1,830 (49.0) |  |
| Statin use -- no.(%) |  |  |  |  |  |  |
| Yes | 179 (41.9) | 1,735 (43.3) | 0.585 | 401 (50.5) | 1,635 (43.8) | <0.001** |
| No | 248 (58.1) | 2,272 (56.7) |  | 393 (49.5) | 2,098 (56.2) |  |

Significance level: \*\*p<0.01; \*p<0.05

### Statistical Analysis

Statistical analysis was conducted using an intention-to-treat approach. We used the same follow-up strategy as SPRINT: patient visits occurred monthly for the first 3 months and then every 3 months thereafter. Follow-up time was censored on the date of the outcome ascertainment^27^. In both intensive and standard arms, baseline characteristics were compared between de-escalation and control patients using the χ^2^ test (or Fisher’s exact test if applicable) for categorical variables and t test for continuous variables. Tests were all 2-sided, and P<0.05 was considered statistically significant. The trends of BP and antihypertensive medication number were described with the mean and standard deviation (SD) of each group at each routine visit. Cox proportional-hazards regressions were applied to estimate the unadjusted and adjusted hazard ratio (HR) of outcomes, incorporating baseline confounders (eTable 1), and Kaplan-Meier curves were used to better visualize the results. We checked treatment changes from screening visit to randomization and constructed sunburst figures visualizing the antihypertensive treatment patterns of each group.

In the sensitivity analysis, we compared health outcomes of intensive (SBP target: <120 mmHg) vs. standard treatment (SBP target: <140 mmHg) in de-escalation patients. To address confounding by indication, we conducted 1:1 propensity score matching with a caliper of 0.25 between de-escalation patients who received intensive treatment and those received standard treatment. Cox proportional-hazards regressions were used to assess outcomes, as in the primary analysis.

## Results

### Baseline characteristics

A total of 8,961 patients were included, with 4,434 and 4,527 patients in the intensive and standard treatment arms, respectively. In the two treatment arms, 427 (9.6%) and 794 (17.5%) patients, respectively, had antihypertensive de-escalation. At randomization, in both the intensive and standard treatment arms, the mean SBP and DBP of de-escalation patients were significantly lower than control patients (Table 1). In the intensive arm, the mean SBP was 134.3 ± 15.6 mmHg for de-escalation patients and 140.0 ± 15.8 mmHg for controls (p<0.001); in the standard arm, the corresponding values were 132.6 ± 15.6 and 140.8 ± 15.8 mmHg (p<0.001). The mean DBP in the intensive arm was 75.5 ± 11.2 mmHg for de-escalation patients and 78.2 ± 12.0 mmHg for controls (p<0.001), while in the standard arm it was 74.4 ± 11.1 and 78.6 ± 12.0 mmHg, respectively (p<0.001) (Table 1).

In the intensive treatment arm, other baseline characteristics of de-escalation and control patients were similar. However, in the standard treatment arm, de-escalation patients were older than controls (mean ± SD age: 68.7 ± 9.4 vs. 67.7 ± 9.5, p=0.007). Besides, a significantly higher proportion of de-escalation patients had moderately to severely impaired kidney (eGFR <60 mL/min/1.73m^2^) function (34.1% vs. 24.9%, p=0.001), received aspirin (56.7% vs. 51.0%, p=0.004), or statin (50.5% vs. 43.8%, p<0.001) than controls.

### Trends in BP and antihypertensive medications

After randomization, in the intensive treatment arm, the mean SBP of de-escalation and control patients dropped quickly from 134.3 and 140.0 mmHg, respectively, to approximately 120 mmHg and remained around 120 mmHg thereafter (Figure 1A). In the standard treatment arm, the mean SBP in de-escalation patients increased from 132.6 to around 135 mmHg and remained stable, while in controls it decreased from 140.8 to around 135 mmHg and plateaued. The DBP in the four groups had qualitatively similar trends as SBP during follow-up (Figure 1A).

**Figure 1.**
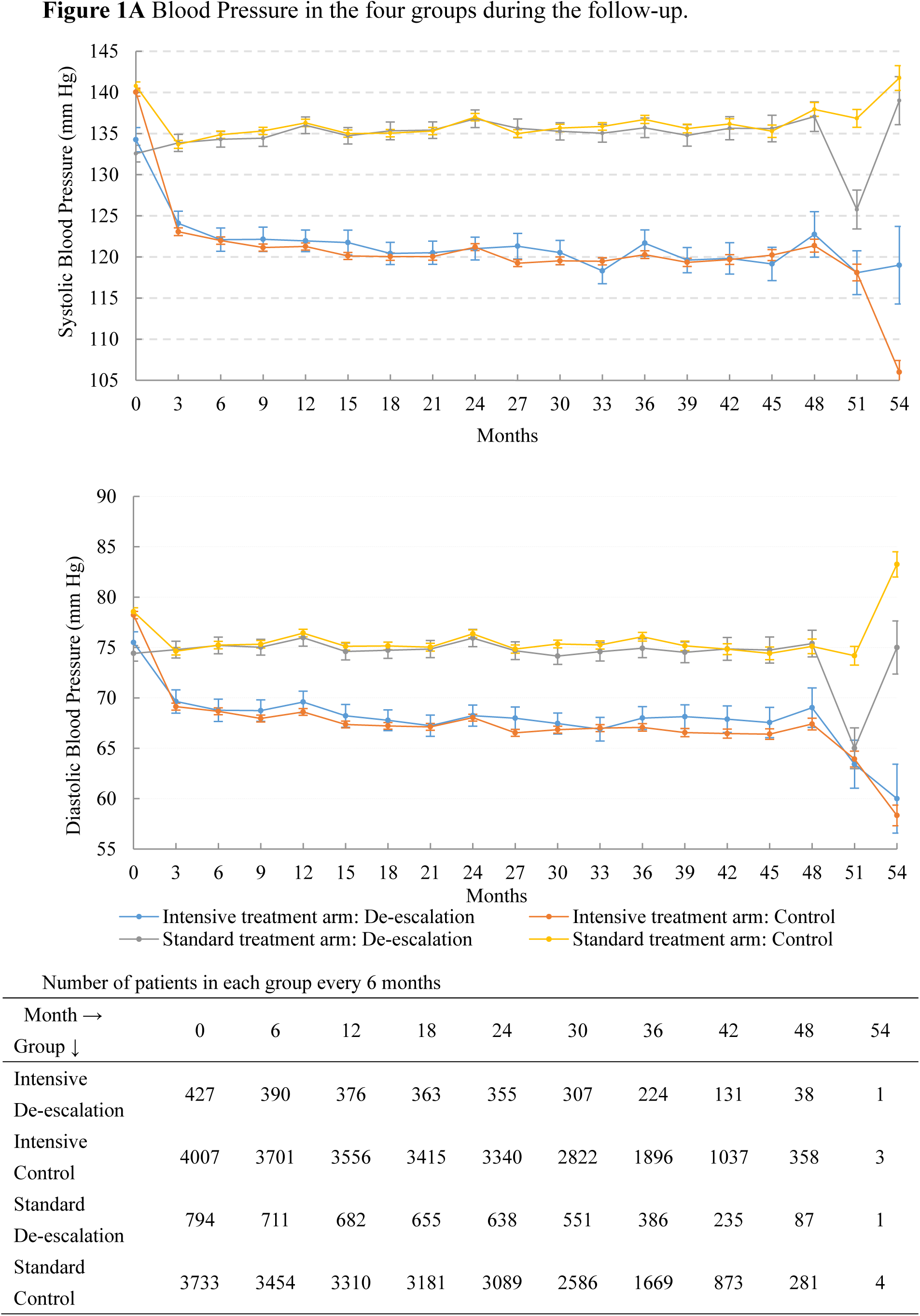
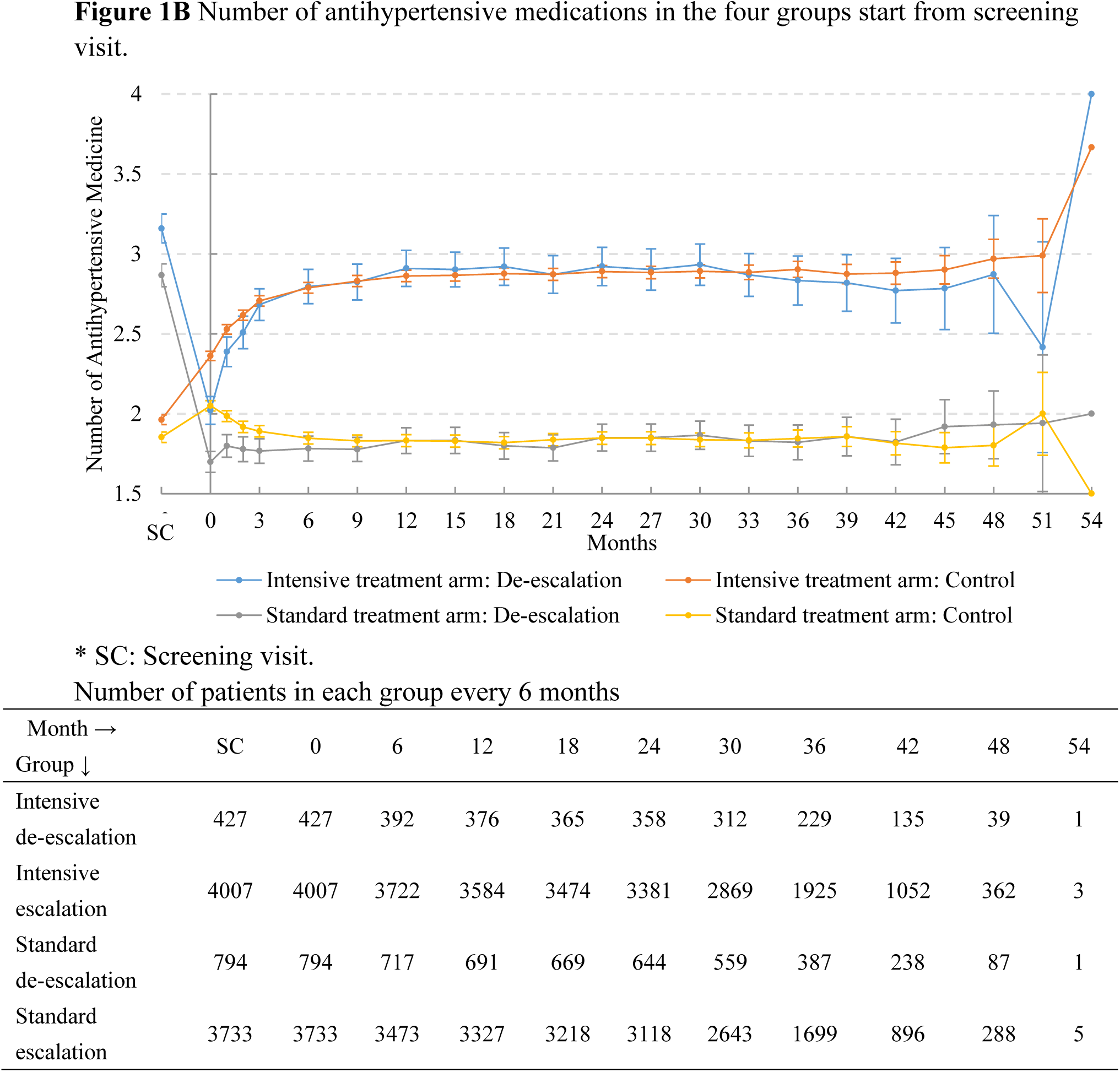
Trends in blood pressure and number of antihypertensive medications

The mean number of medications used are displayed in Figure 1B. Briefly, in the intensive treatment arm, the mean number of medications of de-escalated patients decreased from 3.2 at screening visit to 2.0 at randomization, increasing to around 2.8 from the 12-month visit onward, while for control patients, it increased from 1.96 at screening visit to 2.4 at randomization and around 2.8 by the 12-month visit. In the standard treatment arm, the mean number of medications of de-escalation patients decreased from 2.9 to 1.7 at randomization, increasing to around 1.8, but never reaching pre-randomization level during follow-up. In control patients, it increased from 1.8 to 2.1 at randomization then returned to around 1.8 by month 12 and remained stable. From the screening visit to randomization, ARA, potassium-sparing diuretics, alpha blockers, loop diuretics, vasodilators, and direct renin inhibitor (eTable 2, eFigure 1) were more likely to be de-escalated.

### Outcomes and adverse events

In the intensive treatment arm, the primary outcome occurred on 28 of 427 de-escalation patients (Incidence rate (IR) = 2.07/100 person-year) and 230 of 4,007 control patients (IR = 1.88/100 person-year) (Table 2). The crude HR comparing de-escalation to controls was 1.10 (95% CI: 0.74–1.63; Figure 2A); the adjusted HR was 1.12 (95% CI: 0.75–1.68; Figure 2B), indicating no increased risk after baseline adjustment. For secondary outcomes, de-escalated patients had a 2.47-fold higher incidence of heart failure than controls (Table 2), with an adjusted HR of 2.66 (95% CI: 1.39–5.09; Figure 2B). No significant differences were observed for other secondary outcomes or adverse events in the intensive arm (Figure 2A–B). Kaplan-Meier curve also showed an increased risk of heart failure among de-escalation patients (Figure 3A), and no significant differences between de-escalation and control patients for other outcomes (eFigure 2).

**Figure 2.**
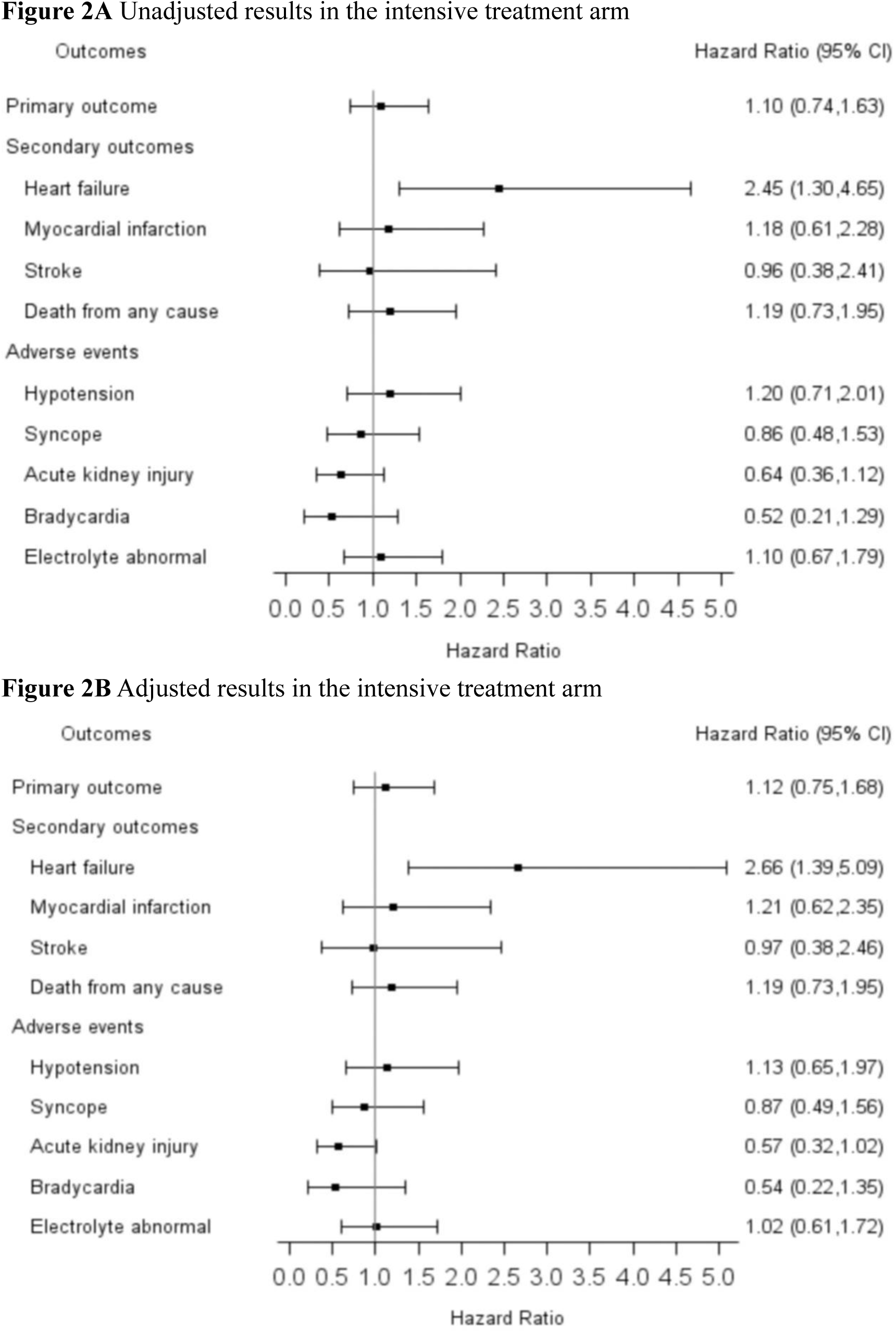
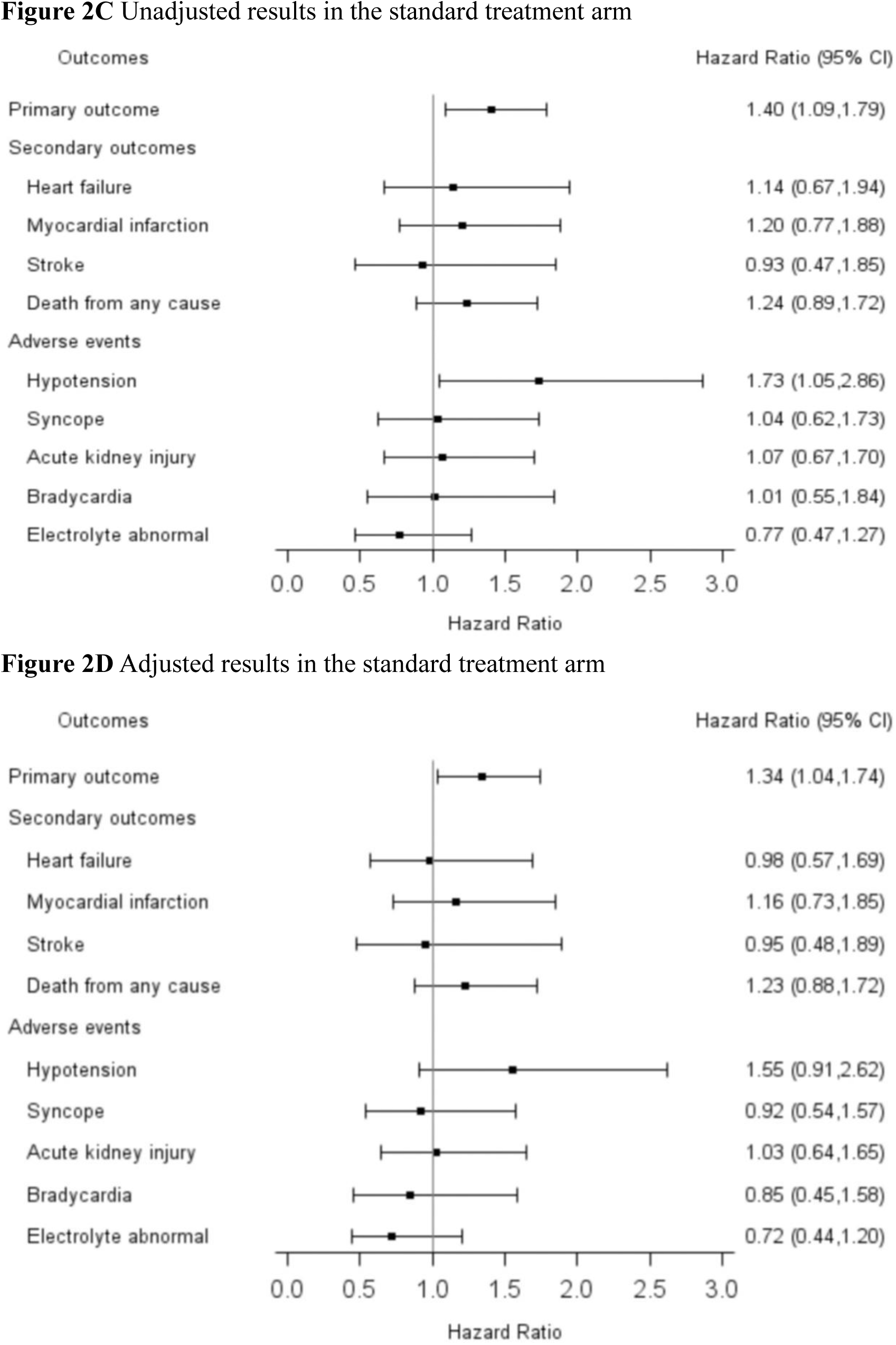
Forest plots for outcomes and adverse events in the intensive and standard treatment arms (hazard ratios represent de-escalation relative to control (ref))

**Figure 3.**
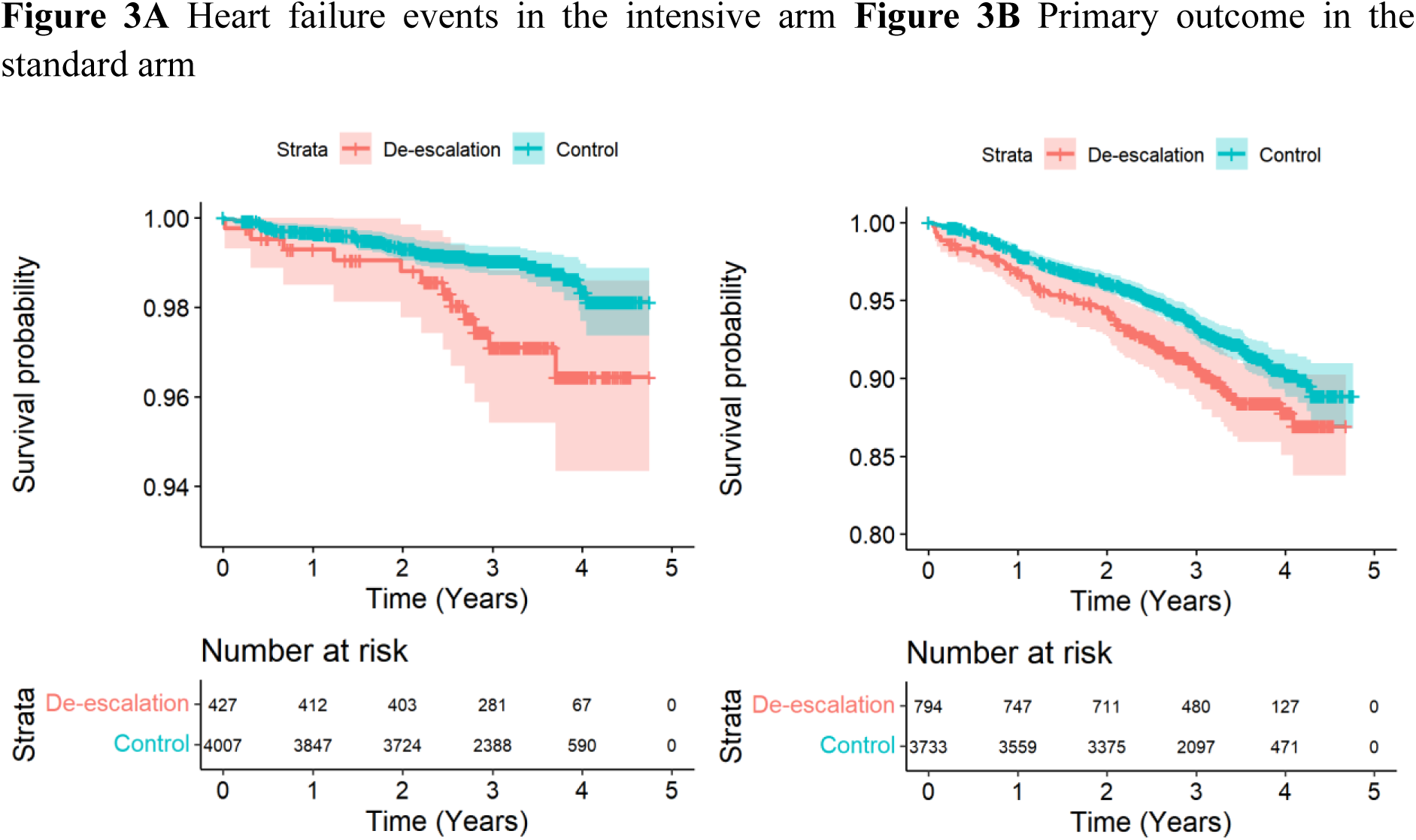
Kaplan-Meier curves for outcomes in the intensive and standard treatment arms

**Table 2.** Number and incidence of effectiveness and safety outcomes (by treatment arm)

| Outcomes | Treatment de-escalation |  |  | Control |  |  | IRR (95% CI) |
| --- | --- | --- | --- | --- | --- | --- | --- |
|  | n | Exposure time<br>(person-year) | Incidence rate<br>(per 100<br>person-year) | n | Exposure time<br>(person-year) | Incidence rate<br>(per 100<br>person-year) |  |
| **Intensive treatment arm** | N=427 |  |  | N=4,007 |  |  |  |
| Primary outcome | 28 | 1,350.13 | 2.07 | 230 | 12,238.23 | 1.88 | 1.10 (0.74 - 1.63) |
| Secondary outcomes |  |  |  |  |  |  |  |
| Heart failure | 12 | 1,374.32 | 0.87 | 44 | 12,455.61 | 0.35 | 2.47 (1.30 - 4.68) |
| Myocardial infarction | 10 | 1,369.80 | 0.73 | 77 | 12,388.67 | 0.62 | 1.17 (0.61 - 2.26) |
| Stroke | 5 | 1,379.42 | 0.36 | 47 | 12,445.14 | 0.38 | 0.96 (0.38 - 2.43) |
| Death from any cause | 18 | 1,394.70 | 1.29 | 134 | 12,560.23 | 1.07 | 1.21 (0.74 - 1.98) |
| Adverse events |  |  |  |  |  |  |  |
| Hypotension | 16 | 1,366.71 | 1.17 | 121 | 12,306.09 | 0.98 | 1.19 (0.71 - 2.00) |
| Syncope | 13 | 1,372.43 | 0.95 | 129 | 12,290.30 | 1.05 | 0.90 (0.51 - 1.59) |
| Acute kidney injury | 13 | 1,374.28 | 0.95 | 181 | 12,248.89 | 1.48 | 0.64 (0.36 - 1.12) |
| Bradycardia | 5 | 1,376.36 | 0.36 | 86 | 12,382.15 | 0.70 | 0.52 (0.21 - 1.28) |
| Electrolyte abnormal | 18 | 1,356.92 | 1.33 | 149 | 12,254.26 | 1.22 | 1.09 (0.67 - 1.78) |
| **Standard treatment arm** | N=794 |  |  | N=3,733 |  |  |  |
| Primary outcome | 80 | 2,429.36 | 3.29 | 264 | 11,277.01 | 2.34 | 1.41 (1.10 - 1.81) |
| Secondary outcomes |  |  |  |  |  |  |  |
| Heart failure | 17 | 2,492.01 | 0.68 | 68 | 11,489.94 | 0.59 | 1.15 (0.68 - 1.96) |
| Myocardial infarction | 24 | 2,484.77 | 0.97 | 92 | 11,441.37 | 0.80 | 1.20 (0.77 - 1.88) |
| Stroke | 10 | 2,513.49 | 0.40 | 49 | 11,494.93 | 0.43 | 0.93 (0.47 - 1.84) |
| Death from any cause | 45 | 2,535.66 | 1.78 | 164 | 11,627.84 | 1.41 | 1.26 (0.91 - 1.75) |
| Adverse events |  |  |  |  |  |  |  |
| Hypotension | 21 | 2,491.97 | 0.84 | 56 | 11,478.67 | 0.49 | 1.73 (1.05 - 2.86) |
| Syncope | 18 | 2,492.80 | 0.72 | 79 | 11,474.11 | 0.69 | 1.05 (0.63 - 1.75) |
| Acute kidney injury | 22 | 2,498.90 | 0.88 | 94 | 11,458.28 | 0.82 | 1.07 (0.67 - 1.71) |
| Bradycardia | 13 | 2,500.37 | 0.52 | 59 | 11,493.25 | 0.51 | 1.01 (0.41 - 2.52) |
| Electrolyte abnormal | 18 | 2,488.20 | 0.72 | 106 | 11,432.64 | 0.93 | 0.78 (0.47 - 1.29) |

In the standard treatment arm, 80 out of 794 de-escalation patients (IR = 3.29/100 person-year) and 264 out of 3,733 control patients (IR = 2.34/100 person-year) developed the primary outcome (Table 2). The unadjusted HR was 1.40 (95% CI: 1.09–1.79; Figure 2C), and the adjusted HR was 1.34 (95% CI: 1.04–1.74; Figure 2D), indicating a 34% increased risk with de-escalation. Kaplan–Meier curves supported this finding (Figure 2B). No significant differences were observed for secondary outcomes or adverse events (Figure 2D; eFigure 3).

Among de-escalation patients, after 1:1 propensity score matching, there were 412 patients in each of the intensive and standard treatment arms. The distributions of the propensity scores between the two arms showed substantial overlap (eFigure 4). Compared with standard treatment, patients who received intensive treatment had lower incidence rate of MACE (2.07 vs. 2.75/100 person-year, eTable 3) but showed no significantly difference in Cox regression model (HR: 0.76, 95% CI: 0.46-1.27, eFigure 5).

## Discussion

Benefiting from the unique setting in SPRINT (de-escalation to meet the eligibility criterion before randomization), our findings quantify the harms of de-escalation due to non-clinical factors and highlight the value of lower target SBP for certain patients. We found that de-escalation is associated with a 34% increased risk of MACE among patients who were targeting SBP to <140 mmHg during a median 3.3-year follow-up. Conversely, under an intensive treatment goal to SBP <120 mm Hg, no excess risk of overall MACE was observed with early de-escalation. Patients with treatment de-escalation had higher risk of CVD than controls at randomization: more likely to have kidney disease, using aspirin or statin, which partially explained the intensive target SBP before randomization.

De-escalation patients in both intensive and standard treatment arms received less antihypertensive medications to increase their SBP to 130-180 mmHg (eligibility criterion for entering SPRINT) before randomization. However, the duration from de-escalation to restoration of the same number of antihypertensive medications as they received at screening visit was different between two arms, which contributed to the increased risk of MACE. After randomization, de-escalation patients in standard treatment arm received fewer antihypertensive medications throughout follow-up than at the screening visit (Figure 1B). Thus, their mean SBP increased and remained higher during the follow-up than at the randomization (Figure 1A). Though patients in both de-escalation and control groups shared comparable SBP and DBP after the 3-month visit, de-escalation patients appeared to have lost the protective effect of lower target SBP and were treated sub-optimally during the follow-up. In contrast, de-escalation patients in the intensive treatment arm may have been sub-optimally treated for only a year (indicated by the number of antihypertensive medications received between screening visit and month 12), leading to comparable risk of MACE between de-escalation and control patients. Therefore, treatment de-escalation with following prolonged low-intensity treatment could increase the risk of MACE, and targeting SBP <120 mmHg after de-escalation could be beneficial than <140 mmHg to this patient population. Our sensitivity analysis in de-escalation patients also showed that intensive treatment is associated with a relatively lower rate of MACE than standard treatment (HR: 0.76, 95% CI: 0.46-1.27), although the small sample size attenuated the association toward the null.

In our findings, de-escalation patients showed a higher risk of CVD than control patients at randomization, which was indicated by higher proportion of moderately to severely impaired renal function patients, and higher proportion of patients using aspirin or statins. Previous studies showed that patients with chronic kidney disease have a higher risk of CVD due to a systemic, chronic proinflammatory state^28^. Anti-inflammatory effect of aspirin and the antilipidemic effect of statin are beneficial for CVD prevention^29,30^, thus patients who received these drugs may have underlying conditions with an elevated risk of CVD. Therefore, de-escalated patients may have been treated to a lower SBP than controls due to elevated cardiovascular risk. Indeed, both existing studies and guidelines demonstrated the benefits of lower SBP target for patients with CKD or other conditions. For example, 2021 WHO hypertension guideline recommends an SBP target <140 mmHg for patients without comorbidities and <130 mmHg for those with CKD^9^. Our findings suggest that in SPRINT, physicians had been treating some higher CVD risk patients to a lower SBP value before entering the trial: de-escalation patients may have been treated to an SBP <130mmHg before entering the SPRINT (because they received treatment de-escalation to raise SBP into 130– 180 mmHg), even though this practice has not reached universal agreement nor recommendation by guidelines before 2017 in the US^31^. This reflects physicians’ prudent clinical judgment, further supported by the observation that the most frequently de-escalated medications were not first-line therapies.

While we acknowledge that de-escalation patients exhibited some markers of higher baseline cardiovascular risk, overall, the de-escalation and control patients were largely similar in baseline characteristics reported, and we implemented a comprehensive approach to address confounders. We controlled for cardiovascular risk, including covariates for DBP, BMI, smoking status, eGFR, and relevant comorbidities, in all of our cox regression models. To further adjust for confounders, we matched de-escalation patients in the intensive and standard arm based on the propensity score in our sensitivity analysis.

It is possible that de-escalated patients in the intensive arm had suboptimal ejection fraction, COPD or substance abuse at baseline, contributing to our finding, higher risk of heart failure^32,33^. However, as these unmeasured confounders only accounted for approximately 5% of the observed increase^32^, and the association remained non-significant in the standard arm (despite a doubled sample size), the higher risk are likely driven by limited sample size and lack of access to appropriate medication (i.e. aldosterone receptor antagonists, or loop diuretics) rather than unmeasured confounding.

Our study has several strengths. First, the unique scenario in SPRINT ensures that the antihypertensive treatment de-escalation in our study is protocol-driven to meet eligibility criteria, not influenced by clinical judgment, which helps reduce confounding by indication. Second, as a clinical trial, patients’ lab values were closely monitored and outcomes were adjudicated, both of which reduce probability of misclassification of important study variables.

There are also some limitations in our study. First, BP at the screening visit was unavailable. Instead, we inferred SBP range based on antihypertensive use (e.g., de-escalation patients’ SBP should be < 130 mmHg, because they received treatment de-escalation to increase SBP into 130-180 mmHg at randomization to meet the inclusion criteria). Second, we defined de-escalation based on the reduction in the number of antihypertensive medications without considering the reduction in dose. We adopted this approach to comply with the protocol of SPRINT for defining drug reduction^34^, and to prevent uncertainty over dose reduction, which are often linked to adverse events. Lastly, SBP in SPRINT was measured with varying levels of staff attendance, potentially introducing white coat effects and measurement variation. However, prior post-hoc analyses of SPRINT showed that BP levels were similar whether measured by attended or unattended techniques^35^.

## Conclusions

De-escalation without clinical consideration is associated with an increased risk of major cardiovascular events, especially among patients who were targeted at standard SBP (<140 mm Hg). Our findings reassure the importance of intensive control of SBP, especially among patients with certain comorbidities, such as CKD.

## Data Availability

Not available.

